# Lanreotide for Advanced Pheochromocytoma and Paraganglioma: Results of a Multicenter Phase II Trial

**DOI:** 10.64898/2026.10.01.26364504

**Authors:** Jaydira Del Rivero, Bahar Laderian, Mengxi Zhou, Lyndon Luk, Susan Bates, Tito Fojo

**Author notes:** **Corresponding author: Tito Fojo MD, PhD,** Columbia University Medical Center / Herbert Irving Comprehensive Cancer Center 630 West 168th Street, New York, NY 10032, USA.

## Abstract

**Background:** Pheochromocytomas and paragangliomas (PPGLs) are rare neuroendocrine neoplasms originating from chromaffin cells that express somatostatin receptors (SSTRs) and may therefore be susceptible to somatostatin analogue therapy. Despite this biological rationale, prospective evidence supporting the antiproliferative activity of somatostatin analogues in PPGL is limited. We conducted a multicenter phase 2 clinical trial to assess the efficacy and safety of lanreotide in patients with advanced or metastatic PPGL. [**LAMPARA, NCT03946527**]

**Methods:** Patients with advanced or metastatic PPGL and evidence of recent disease progression received lanreotide depot/autogel 120 mg subcutaneously every 4 weeks. Treatment was planned for 52 weeks with an option to continue for an additional 52 weeks. Endpoints included overall survival (OS), progression-free survival (PFS), and response according to RECIST. Serum chromogranin A (CgA) was evaluated as an exploratory biomarker. A tumor growth rate analysis was planned to allow comparison with data from the CLARINET trial in gastroenteropancreatic NETs.

**Results:** Eighteen patients (median age 42 years; range 28-77) across three centers participated; 78% carried SDHx mutations. Lanreotide was well tolerated with predominantly grade 1-2 adverse events and no treatment discontinuations due to toxicity. The most common treatment-emergent adverse events were diarrhea (67%), injection-site reaction (44%), constipation (39%), fatigue (39%), and abdominal pain (39%). Serious adverse events were reported in four patients and were assessed as not related or unlikely related to lanreotide. All 18 patients had at least one recorded post-baseline tumor assessment. Two partial responses were recorded during extended follow-up. At approximately 1 year, disease stabilization was observed in 11 of 13 patients (85%) with an available assessment, while two patients (15%) had progressive disease. Biomarker analysis showed substantial variability in serum CgA values. Tumor growth rate analysis revealed a median growth rate of 0.00124/day (tumor doubling time 559 days), comparable in order of magnitude to the rate of 0.00046/day observed in 83 lanreotide-treated patients in CLARINET, and consistent with a median PFS ≥2 years.

**Conclusions:** Lanreotide exhibits promising antiproliferative activity in PPGL, with a favorable safety profile and extended periods of disease stabilization. These findings support the consideration of SSAs as an option in managing SSTR-positive PPGLs, delaying more aggressive treatments until disease progression mandates escalation.

## INTRODUCTION

Pheochromocytomas arise from adrenomedullary chromaffin cells, while paragangliomas are derived from extra-adrenal chromaffin cells of the sympathetic paravertebral ganglia of the thorax, abdomen, and pelvis, or the parasympathetic ganglia along the glossopharyngeal and vagal nerves in the neck and skull base. [1, 2] Although approximately 25% of PPGLs develop metastatic disease, these tumors remain among the most challenging neuroendocrine neoplasms to manage, presenting the dual challenges of hormonal excess and malignant behavior.[1]

The treatment landscape for metastatic PPGL has historically been limited. Cytotoxic chemotherapy with cyclophosphamide, vincristine, and dacarbazine (CVD) has been a mainstay, and radiolabeled therapy with meta-iodobenzylguanidine (MIBG) has provided additional options. [1] More recently, the FIRSTMAPPP trial established sunitinib as the first agent with randomized evidence of anti-tumor efficacy in progressive metastatic PPGL, demonstrating a 12-month PFS rate of 36% versus 19% for placebo, with median PFS values of 8.9 versus 3.6 months. [3] The LITESPARK-015 trial subsequently demonstrated the efficacy of belzutifan, a HIF-2α inhibitor, with an objective response rate of 26%, disease control rate of 85%, and median PFS of 22.3 months, leading to the first FDA approval of an oral therapy for PPGL in May 2025. [4] Axitinib, another VEGF-receptor tyrosine kinase inhibitor, has also shown activity in a single-arm phase 2 trial, achieving a partial response rate of 35.3% and median PFS of 7.9 months. [5] PRRT with lutetium-177 dotatate has demonstrated disease control rates of 63-86% in retrospective and prospective studies, though with notable toxicities including catecholamine release syndrome in up to 17% of patients. [6, 7]

Despite being classified as neuroendocrine tumors, PPGLs were not included in the landmark clinical trials that established the antiproliferative role of SSAs in other neuroendocrine tumors. This exclusion persists despite robust evidence that PPGLs express SSTRs, particularly SSTR2, at levels comparable to other NETs. [8–10] SSTR2 expression is especially prominent in SDHx-mutated tumors, with 91% of SDH-deficient tumors demonstrating moderate or strong SSTR2A staining compared to 49% of SDH-sufficient tumors. [11] SSTR-based PET imaging with ^68^Ga-DOTATATE has become an important and highly sensitive imaging modality for metastatic and SDHx-related PPGLs. [12, 13]

The CLARINET trial established the antiproliferative efficacy of lanreotide in gastroenteropancreatic NETs, demonstrating significantly prolonged PFS compared to placebo (HR 0.47; 95% CI, 0.30–0.73) with estimated 24-month PFS rates of 65.1% versus 33.0%.[14] The PROMID trial similarly demonstrated the antiproliferative effect of octreotide LAR in midgut NETs. [15] These findings together, with the known SSTR expression in PPGLs, provided the rationale for evaluating lanreotide in this population.

Current NCCN guidelines list SSAs (octreotide acetate LAR or lanreotide) as "Other Recommended" options for SSTR-positive PPGL, while noting that data about antiproliferative effects of SSAs in this setting are limited and clinical trials are ongoing. [13] We report the results of a multicenter, single-arm phase 2 trial evaluating the efficacy and safety of lanreotide in patients with advanced or metastatic PPGL and recent disease progression.

## METHODS

### Study Design and Patients

This was a multicenter, single-arm, open-label, phase 2 clinical trial evaluating the efficacy and toxicity of lanreotide depot/autogel in patients with advanced or metastatic pheochromocytoma or paraganglioma. The study was conducted at three sites: Columbia University Irving Medical Center (CUMC), Cleveland Clinic and the National Institutes of Health. Eligible patients were adults with a confirmed diagnosis of PPGL with advanced or metastatic disease. Evidence of recent disease progression while either not receiving any therapy or receiving a therapy deemed ineffective was required for enrollment. Prior therapies were permitted, including surgery, radiation therapy, chemotherapy, and PRRT.

### Treatment

Patients received lanreotide depot/autogel 120 mg administered subcutaneously every 4 weeks. Treatment was planned for 52 weeks (13 cycles) with the option to continue for an additional 52 weeks (total 104 weeks) in patients deriving clinical benefit.

### Endpoints and Assessments

The primary endpoints included OS, PFS, and tumor response according to RECIST criteria. Safety was assessed using the Common Terminology Criteria for Adverse Events (CTCAE). Serum chromogranin A (CgA) was measured serially at protocol-specified visits as a potential biomarker of response.

Given the rarity of these cancers, a novel tumor growth rate analysis was planned to allow comparison with data from the CLARINET trial. This methodology, previously validated in the CLARINET dataset, estimates exponential rate constants for tumor growth (g) and regression/decay (d), with the rate of tumor growth, designated ***g***, serving as a biomarker for overall survival and treatment effectiveness. [16] Tumor growth rates and doubling times were intended to be calculated for evaluable patients and compared with the growth rates observed in 83 lanreotide-treated patients from CLARINET.

### Statistical Analysis

Descriptive statistics were used to summarize patient characteristics, treatment outcomes, and adverse events. PFS was estimated using the Kaplan-Meier method. Tumor growth rates were planned to be compared descriptively with historical data from the CLARINET trial. Adverse-event incidence was calculated using the maximum grade recorded for each patient and adverse-event term.

## RESULTS

### Patient Characteristics

Eighteen patients were enrolled with a median age of 42 years (range 28-77). The cohort included 11 females (61%) and 7 males (39%). Self-reported race included 13 White patients (72%), two Black or African American patients (11%), two patients of another race (11%), and one patient with unknown race (6%). Ethnicity was reported as not Hispanic/Latino in 10 patients (56%), other Hispanic in two (11%), and unknown in six (33%). Notably, 14 of 18 patients (78%) harbored an SDHx germline mutation, consistent with the known association between SDHx mutations and metastatic PPGL.[8, 11]

Sixteen of 18 patients (89%) had undergone prior surgery, with procedures including adrenalectomy, retroperitoneal paraganglioma resection, partial cystectomy, carotid body tumor resection, hepatic metastasectomy, lymph node dissection, and lung biopsy. Several patients had undergone multiple surgical procedures (range 1-3 operations). Prior systemic and radiation therapies were documented in a subset of patients: radiation therapy was administered in 5 patients, including three-dimensional conformal radiation therapy, stereotactic body radiation therapy (SBRT), proton beam radiation, and MIBG therapy. Prior systemic therapies included CVD chemotherapy (1 patient), axitinib (1 patient), and mFOLFIRINOX followed by gemcitabine-abraxane (1 patient, initially treated for a concurrent diagnosis). No patient had received prior SSA therapy. Diagnosis dates ranged from 1997 to 2021, with time from initial diagnosis to study enrollment ranging from approximately 1 to 26 years, reflecting the heterogeneous and often protracted clinical course of metastatic PPGL **[Table 1].**

**Table 1.** Baseline Patient Characteristics.

| **Table 1 - Baseline Patient Characteristics** |  |  |
| --- | --- | --- |
| **Characteristic** |  | **N = 18** |
| Age, median (range), years |  | 42 (28–77) |
| Sex |  |  |
|  | Female, n (%) | 11 (61) |
|  | Male, n (%) | 7 (39) |
| Race |  |  |
|  | White, n (%) | 13 (72) |
|  | Black or African American, n (%) | 2 (11) |
|  | Other, n (%) | 2 (11) |
|  | Unknown, n (%) | 1 (6) |
| SDHx mutation, n (%) |  | 14 (78) |
| Metastatic disease, n (%) |  | 18 (100) |
| Prior therapies |  |  |
|  | Surgery, n (%) | 16 (89) |
|  | Radiation therapy, n (%) | 5 (28) |
|  | Systemic therapy, n (%) | 3 (17) |
| ECOG Performance Status |  |  |
|  | ECOG PS 0, n (%) | 16 (89) |
|  | ECOG PS 1, n (%) | 2 (11) |

### Efficacy

All 18 patients had at least one recorded post-baseline overall tumor assessment. Two patients (11%) had a partial response recorded, 15 patients (83%) had stable disease as their best recorded response including five who eventually had progression of their disease, and one patient (6%) had progressive disease as the best recorded response. [**Figure 1**] Reductions in metanephrines, normetanephrines and chromogranin were observed in a similar fraction of patients [**Table 2**] The median PFS had not been reached and exceeded 2 years. [**Figure 2**] Two patients (11%) who discontinued study participation at one year due to the burden of traveling were censored at one year.

**Figure 1.**
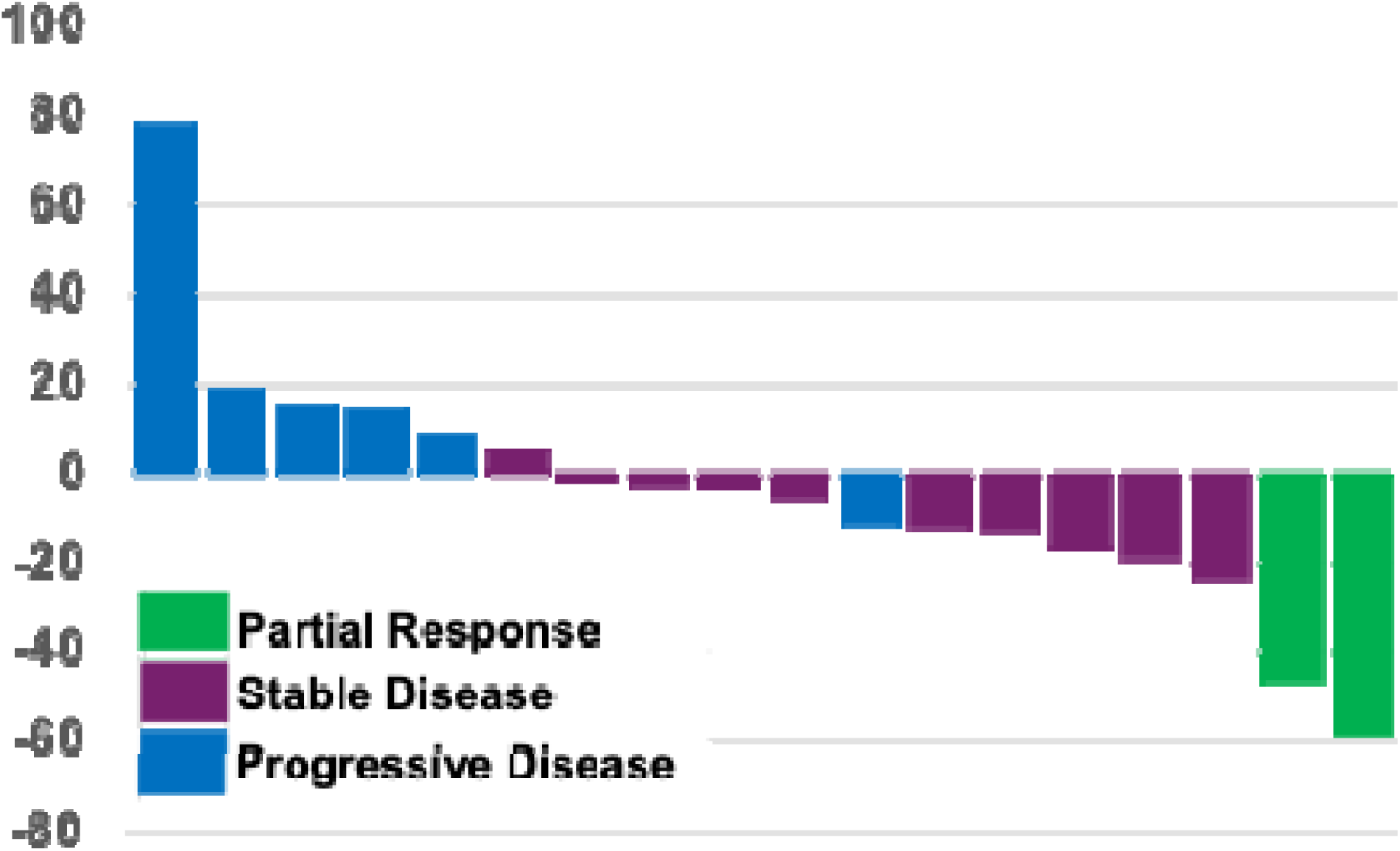
Waterfall Plot. Colors indicate the final assessment while on study for 1’-2 years.

**Figure 2.**
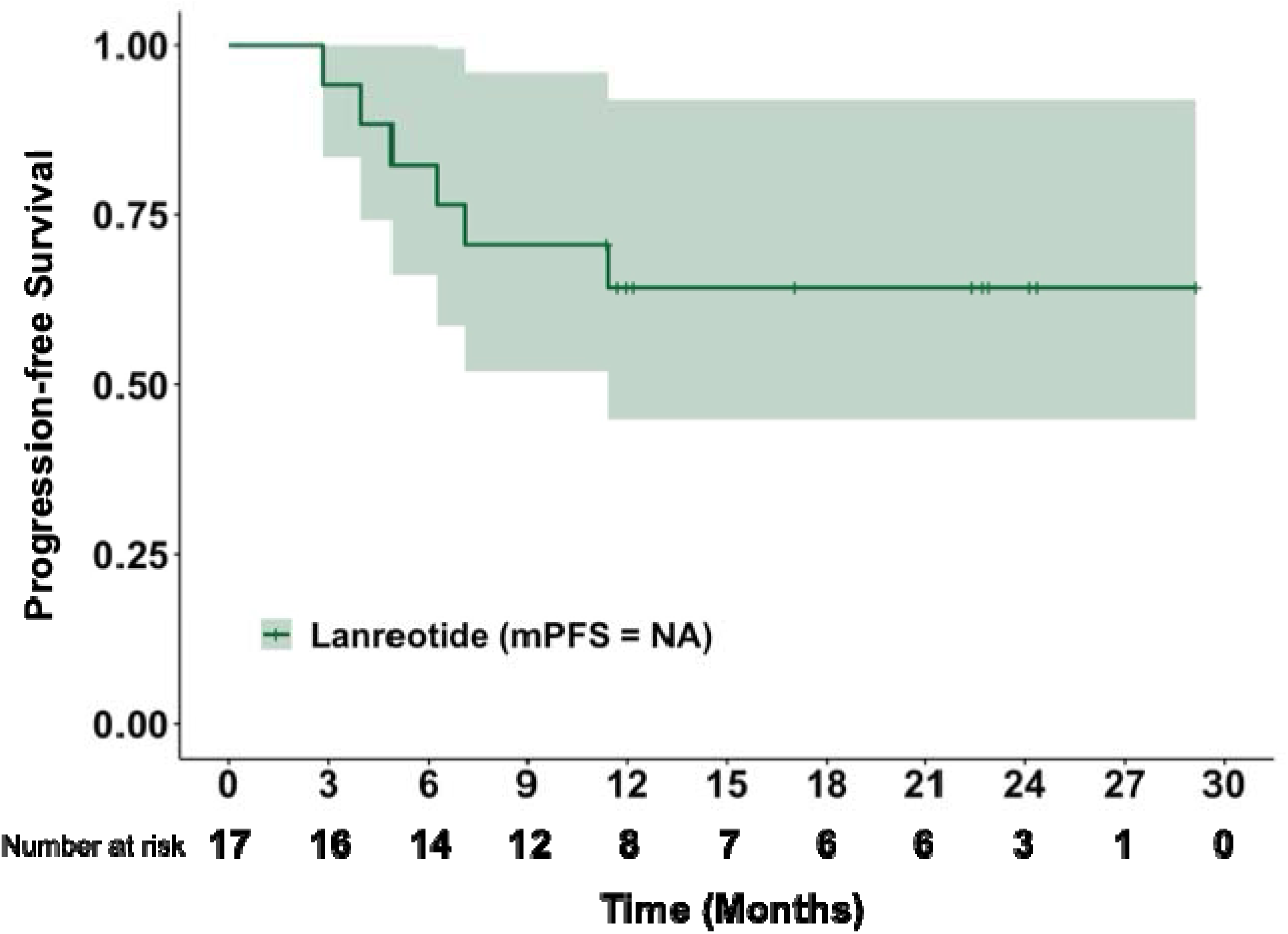
Kaplan Meier plot of Progression-free survival. The median PFS was not reached and exceeds 2 years. Vertical bars indicate censoring times. Two bars close to 12 months describe two patients who for personal reasons discontinued trial participation after the end of the first year and chose not to continue onto the extension phase

**Table 2.** Biochemical Response. [Definition: >20% drop in 24-hour urinary analysis, sustained for >12 weeks]

|  |  |
| --- | --- |
| Metanephrine Response (*n, %*) |  |
| Yes | 5 (38) |
| No | 7 (54) |
| Unknown | 1 (8) |
| Normetanephrine Response (*n, %*) |  |
| Yes | 3 (23) |
| No | 9 (69) |
| Unknown | 1 (8) |
| Chromogranin A Response (*n, %*) |  |
| Yes | 5 (38) |
| No | 7 (54) |
| Unknown | 1 (8) |

### Chromogranin A Biomarker Analysis

Serum CgA was measured serially in all 18 patients, although collection was not performed at every protocol-specified visit. CgA values varied widely across the cohort. Each patient had at least three numeric CgA measurements, with recorded values ranging from 15 to 169,600 in the reported units. Four patients had markedly elevated recorded CgA values: patient 001-001 had values ranging from 8,480 to 13,560 ng/mL; patient 001-003 had values ranging from 12,120 to 26,010 ng/mL; patient 001-010 had values ranging from 506 to 2,559 ng/mL; and patient S2820-002-02 had values ranging from 23,320 to 169,600 ng/mL. Patient S2820-003-05 demonstrated a rising CgA trajectory from 146 to 1,008 ng/mL over approximately 12 months, which coincided with subsequent disease progression. Reference values are not universal because different diagnostic kits use different antibodies and calibration techniques, but upper limits of normal range were 100-187. Overall, CgA values demonstrated substantial interpatient variability and did not show a consistent relationship with radiographic disease status.

### Tumor Growth Rate Analysis

Rates of tumor growth and regression could be assessed in 16 of 18 patients (89%). Growth was not detected in 4 of 16 evaluable patients (25%) [**Figure 3**]. In the remaining 11 patients with estimable growth, the median tumor growth rate was 0.00124/day, corresponding to a tumor-doubling time of 559 days. This compares with a median growth rate of 0.00046/day [doubling time 1506 days] observed in 83 patients treated with lanreotide in the CLARINET trial. [16] While the growth rate in the PC/PG cohort was numerically higher than in the CLARINET population, the comparable magnitude suggests a similar degree of growth retardation by lanreotide across these tumor types.

**Figure 3.**
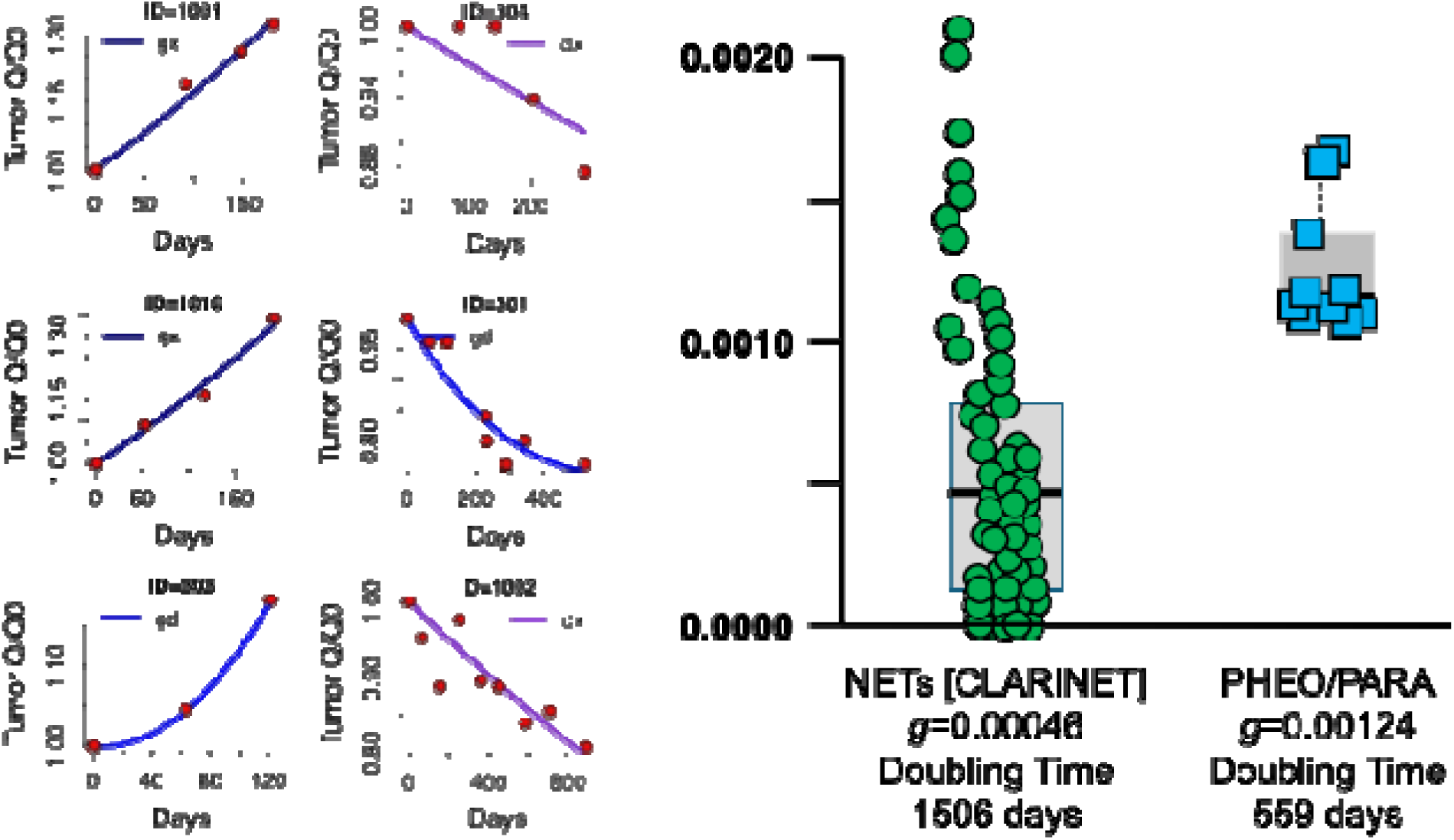
Fit plots on the left show the results in six patients including three on the left who had progressive disease and three on the right of patients whose disease was scored as stable disease having reductions in the sum of the maximum diameters less than 30%. The red dots are the data obtained by imaging. The black line has been drawn by the ***tumgr*** program as provides the best fit that reduced the Akaike Information Criterion (AIC). ***gx*** describes a tumor where only growth could be detected; ***dx*** a tumor in which only regression was observed; ***gd*** a tumor in which both growth and regression could be discerned to be occurring simultaneously. On the right one can see a comparison of the growth rates of gastroenteropancreatic neuroendocrine tumors treated with lanreotide in the CLARINET trial. This is compared with the growth rate of pheochromocytomas and paragangliomas in this, the LAMPARA trial. ***tumgr program*** - https://wilkersj.shinyapps.io/tumgrShiny, http://wilkersj.github.io/tumgr, https://github.com/wilkersj/tumgr

### Safety

Lanreotide was well tolerated. Among 228 treatment-emergent adverse-event records, 165 (72%) were grade 1, 45 (20%) were grade 2, 16 (7%) were grade 3, one (<1%) was grade 4, and one (<1%) was grade 5. No unexpected toxicities were observed. Importantly, no patient discontinued treatment due to adverse events. Blood pressure remained clinically stable throughout treatment, and no consistent pattern of clinically significant ECG, hematologic, or serum chemistry abnormalities attributable to lanreotide was observed **(Table 3).** Adverse events were classified by the maximum grade recorded for each patient and included only events beginning after initiation of lanreotide; baseline and pre-existing conditions were excluded.

**Table 3.**
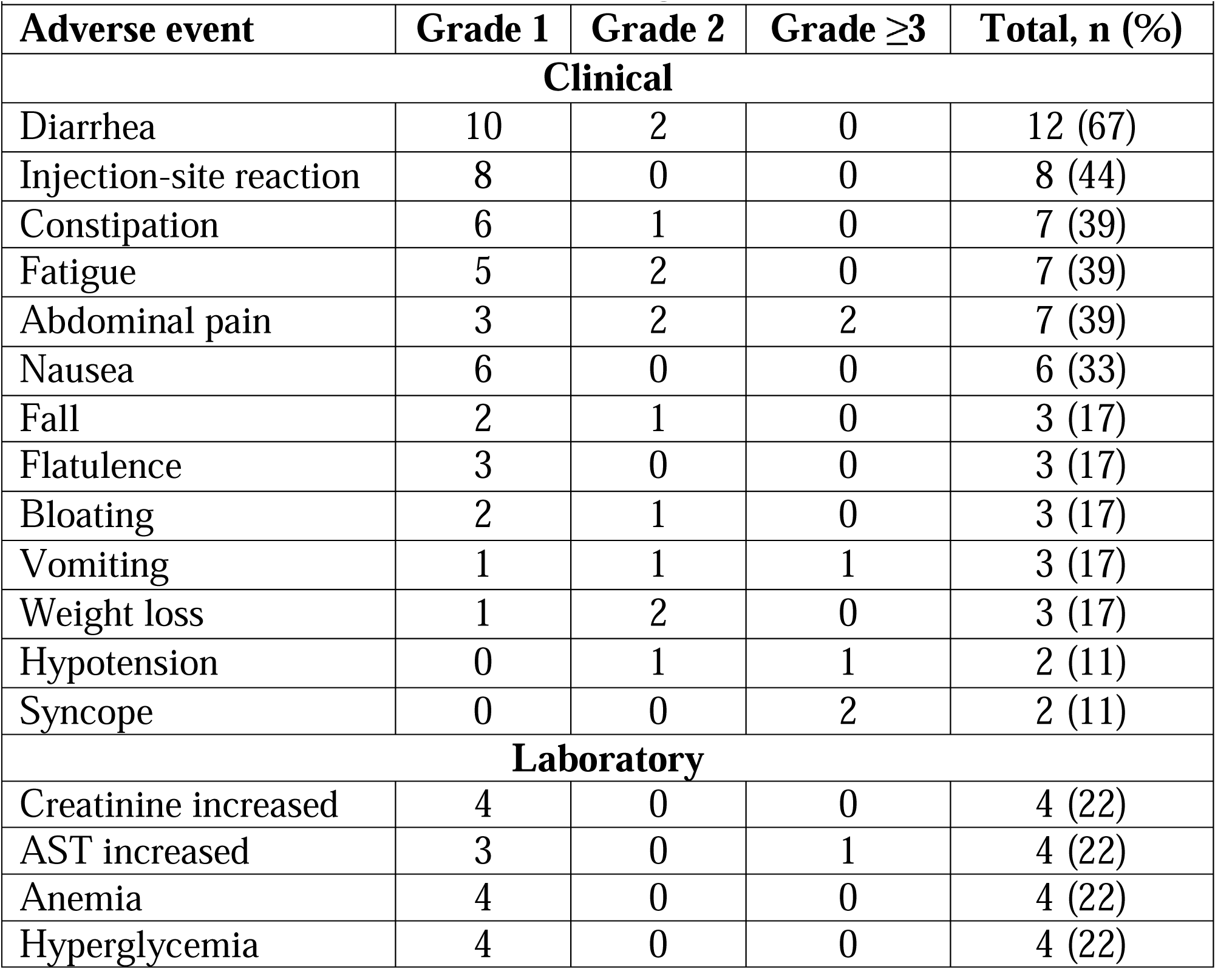
Treatment-Emergent Adverse Events.

| **Table 3 - Treatment-Emergent Adverse Events** |  |  |  |  |
| --- | --- | --- | --- | --- |
| **Adverse event** | **Grade 1** | **Grade 2** | **Grade $\geq 3$** | **Total, n (%)** |
| **Clinical** |  |  |  |  |
| Diarrhea | 10 | 2 | 0 | 12 (67) |
| Injection-site reaction | 8 | 0 | 0 | 8 (44) |
| Constipation | 6 | 1 | 0 | 7 (39) |
| Fatigue | 5 | 2 | 0 | 7 (39) |
| Abdominal pain | 3 | 2 | 2 | 7 (39) |
| Nausea | 6 | 0 | 0 | 6 (33) |
| Fall | 2 | 1 | 0 | 3 (17) |
| Flatulence | 3 | 0 | 0 | 3 (17) |
| Bloating | 2 | 1 | 0 | 3 (17) |
| Vomiting | 1 | 1 | 1 | 3 (17) |
| Weight loss | 1 | 2 | 0 | 3 (17) |
| Hypotension | 0 | 1 | 1 | 2 (11) |
| Syncope | 0 | 0 | 2 | 2 (11) |
| **Laboratory** |  |  |  |  |
| Creatinine increased | 4 | 0 | 0 | 4 (22) |
| AST increased | 3 | 0 | 1 | 4 (22) |
| Anemia | 4 | 0 | 0 | 4 (22) |
| Hyperglycemia | 4 | 0 | 0 | 4 (22) |

Serious adverse events were reported in four of 18 patients (22%). Patient 001-003 experienced grade 3 confusion, grade 3 syncope, grade 3 small-intestinal obstruction, grade 3 left-ventricular systolic dysfunction, grade 4 pericardial tamponade, and grade 5 sudden death; all were assessed as not related to lanreotide and attributed to underlying disease. Patient S2820-002-02 experienced grade 3 autonomic dysfunction, grade 3 abdominal pain, grade 3 vomiting, and grade 3 ileus; all were assessed as not related to lanreotide. Patient 001-009 experienced grade 3 syncope, which was assessed as not related to lanreotide. Patient S2820-003-01 experienced a grade 3 lung infection, which was assessed as unlikely related to lanreotide. No serious adverse event was assessed as probably or definitely related to lanreotide.

## DISCUSSION

This phase 2 trial provides prospective evidence regarding the activity and safety of lanreotide in patients with advanced or metastatic pheochromocytoma and paraganglioma. The results demonstrate that lanreotide was generally well tolerated and was associated with prolonged disease stabilization in a substantial proportion of patients all of whom had experience disease progression close to enrollment.

Fifteen of eighteen patients (83%) had stable disease as their best recorded response, two patients (11%) achieved a partial response during extended follow-up, and one patient (6%) had progressive disease as the best recorded response. The overall disease-control rate was 94%. At the 1-year assessment, 12 patients (66%) had stable disease/partial response and six (34%) had progressive disease.

A median PFS exceeding 2 years compares favorably with other systemic therapies evaluated in metastatic PPGL. In the FIRSTMAPPP trial, sunitinib achieved a 12-month PFS rate of 36% with a median PFS of 8.9 months, though this was in a population that might be considered more difficult to treat. [3] The SUTNET trial reported a 12-month PFS rate of 53.4% and median PFS of 14.1 months with sunitinib. [17] In a similar population, axitinib demonstrated a median PFS of 7.9 months [5]. Belzutifan, the most recently approved agent, achieved a median PFS of 22.3 months in the LITESPARK-015 trial, with an objective response rate of 26% and disease control rate of 85%.[4] PRRT with 177Lu-DOTATATE has shown median PFS values ranging from 13 to 26.9 months across studies, with a recent prospective phase 2 trial reporting median PFS of 19.9 months and a 6-month PFS rate of 86.1%.[6, 7, 18]

It is important to note that direct cross-trial comparisons must be interpreted with caution due to differences in patient populations, disease biology, and eligibility criteria. The current study enrolled patients with evidence of recent disease progression, though the rate and extent of progression at baseline may differ from the strictly defined progressive disease required in trials such as FIRSTMAPPP. The high proportion of SDHx-mutated patients (78%) in our cohort may also influence outcomes, as SDHx-related PPGLs tend to have high SSTR2 expression and may be particularly amenable to SSA therapy. Nevertheless, the prolonged PFS observed with lanreotide is noteworthy and suggests meaningful antiproliferative activity.

### Tumor Growth Rate Analysis

The novel tumor growth rate methodology employed in this study provides a quantitative framework for comparing treatment effects across different tumor types and trial populations. [16] The median tumor growth rate of 0.00124/day in PC/PG patients treated with lanreotide, corresponding to a doubling time of 559 days, compares with a rate of 0.00046/day [doubling time 1506 days] observed in CLARINET. Previous analysis of the CLARINET data demonstrated that lanreotide primarily slows tumor growth rather than accelerates tumor regression, and that this growth-retarding effect is sustained over prolonged periods of administration. [16] The tumor-growth findings in the present study are consistent with a prolonged growth-retarding effect in some patients with PPGL; however, the historical comparison does not establish equivalent efficacy between the two tumor populations.

### Chromogranin A as a Biomarker

The limited utility of CgA as a biomarker in this study merits discussion. Although CgA is widely used as a circulating biomarker in gastroenteropancreatic NETs, its role in PPGL is less well established. In this study, serial CgA values varied substantially among patients, ranging from 15 to 169,600 in the recorded units. Four patients had markedly high recorded values, while most patients had substantially lower values throughout treatment. Serial measurements did not demonstrate a consistent relationship with radiographic disease status across the cohort. These findings suggest that CgA should not be used as the sole measure of treatment response in PPGL and that radiographic assessment remains essential for disease monitoring. The prospective ^177^Lu-DOTATATE trial by Lin et al. found that plasma chromogranin A and normetanephrine were the best tumor-marker surrogates in that setting, suggesting that the biomarker landscape may differ depending on the therapeutic modality and patient population.[6]

### Safety Considerations

The updated adverse-event analysis confirms that lanreotide was generally well tolerated in this PPGL population. The most common treatment-emergent adverse events were diarrhea (67%), injection-site reaction (44%), constipation (39%), fatigue (39%), and abdominal pain (39%). These events were predominantly grade 1 or 2 and were consistent with the established safety profile of lanreotide in other neuroendocrine tumor populations. No patient discontinued treatment because of an adverse event. Four patients (22%) experienced serious adverse events. No serious adverse event was assessed as probably or definitely related to lanreotide. The most notable case was patient 001-003, who experienced multiple serious cardiovascular events including pericardial tamponade and ultimately sudden death, all assessed as unrelated to lanreotide and consistent with the known cardiovascular complications of catecholamine-secreting tumors.

This safety profile contrasts with the profile of other systemic therapies used in the treatment of PPGL. Sunitinib was associated with grade 3-4 adverse events in 61% of patients in FIRSTMAPPP, with 14% discontinuing treatment. [3] Axitinib produced grade 3 adverse events in 59% of patients, predominantly hypertension. [5] Belzutifan caused grade 3 anemia in 22% of participants, and treatment-related serious adverse events occurred in 11%. [4] PRRT with 177Lu-DOTATATE carries risks of grade 3+ hematologic toxicity in 13–20% of patients and catecholamine release syndrome in up to 17%, with some patients requiring ICU admission. [6, 7, 19] The uneventful blood pressure control observed with lanreotide is particularly significant. Hypertension management is a major clinical challenge in patients with PPGL, and therapies that exacerbate hypertension, such as TKIs, require careful monitoring and dose adjustments. [1] The absence of blood pressure complications with lanreotide can be seen as clinically advantageous. Notably, belzutifan demonstrated a complementary benefit in this regard, with 32% of participants achieving a ≥50% reduction in antihypertensive medication dose sustained for ≥6 months. [4]

### Somatostatin Receptor Expression and Rationale

The biological rationale for SSA therapy in PPGL is well established. PPGLs express SSTRs, particularly SSTR2, at levels comparable to other NETs. [9, 10] This expression is especially prominent in SDHx-mutated tumors, which comprised 78% of the current study population. [8, 11] SSTR2 positivity has been independently associated with both SDHx mutations and metastatic disease. [8] A retrospective multicenter analysis reported a disease control rate of 100% with first-line cold somatostatin analogs in metastatic PPGLs, though the sample size was small (n = 6). [20] The current study substantially expands the evidence base for SSA therapy in this population.

### Implications for Treatment Sequencing

The current NCCN guidelines list SSAs as "Other Recommended" therapy for SSTR-positive PPGL, while noting that data about antiproliferative effects are limited. [13] The present results provide prospective evidence that lanreotide can produce prolonged disease control with generally manageable toxicity in selected patients with advanced PPGL. These findings suggest that lanreotide may be considered as one treatment option for patients with SSTR-positive disease, particularly when a treatment with relatively low toxicity is clinically appropriate. The favorable efficacy and safety profile of lanreotide supports a treatment sequencing strategy analogous to that employed in gastroenteropancreatic NETs: initiate therapy with an SSA, extract its benefit through prolonged disease stabilization, and delay more toxic therapies such as PRRT until meaningful, consistent disease progression is documented. This approach is supported by the NCCN guidelines for GEP-NETs, where SSAs are recommended as preferred first-line therapy for SSTR-positive tumors, with PRRT reserved for progression on SSA therapy.[13]. However, this single-arm study does not establish lanreotide as the preferred first-line therapy or define its optimal sequence relative to belzutifan, tyrosine-kinase inhibitors, high-specific-activity MIBG, or PRRT.

PRRT with ^177^Lu-DOTATATE can provide clinically meaningful disease control in PPGL but carries risks that include hematologic toxicity and catecholamine release syndrome. [19] A recent prospective phase 2 trial of 177Lu-DOTATATE in PPGL reported a 17% incidence of grade 3+ catecholamine release syndrome, with some patients requiring ICU admission. [6] The median PFS of 19.9 months in that trial, while clinically meaningful, was not substantially longer than the median PFS exceeding 2 years observed with lanreotide in the current study, further supporting the rationale for initial SSA therapy. The approval of belzutifan in May 2025 adds an important new option to the treatment landscape. [4] The optimal sequencing of SSAs, belzutifan, TKIs, and PRRT remains to be defined and will likely depend on individual patient factors including genotype, SSTR expression, disease tempo, and comorbidities. We leave up to practicing physician the decision as to when to administer lanreotide therapy. While in patients with very aggressive disease a therapy such as sunitinib or CVD that can achieve a higher objective response rate might be considered, in patients with more indolent presentations lanreotide might be seen as a useful first therapy.

### Limitations

This study has certain limitations inherent to its design as a single-arm, phase 2 trial with a relatively small sample size. The absence of a control group limits the ability to draw definitive conclusions regarding the treatment effect in comparison to the natural disease course. The high proportion of patients with SDHx mutations (78%) may impact the generalizability of the findings to patients with sporadic PPGL. While the comparison of tumor growth rates with the CLARINET study provides valuable insights, it involves different tumor types, which may have distinct natural histories.

### Conclusions

This phase 2 study demonstrates that lanreotide was generally well tolerated and was associated with prolonged disease control in a substantial proportion of patients with advanced or metastatic PPGL. The objective response rate was 11% and the median PFS longer than 2 years, with no treatment discontinuations attributed to adverse events. These findings support lanreotide as a treatment option for SSTR-positive PPGL. Future studies should explore the optimal sequencing of SSAs with other active agents including belzutifan, TKIs, CVD, and PRRT. Biomarker-driven approaches incorporating SSTR expression levels, SDHx mutation status, and tumor growth rate kinetics may help identify patients most likely to benefit from SSA therapy.

## Data Availability

The data generated and analyzed during the present study are available from the corresponding author upon reasonable request, subject to applicable institutional policies, ethical requirements, and participant privacy protections.

## REFERENCES

1. Glover, M., et al., Approach to the patient with metastatic pheochromocytoma and paraganglioma: advances in systemic therapy. J Clin Endocrinol Metab, 2026. 111(8): p. 2389–2402.

2. Ghosh, I., et al., Update on Systemic Therapies for Metastatic/Unresectable Pheochromocytomas and Paragangliomas and Future Directions. Cancers (Basel), 2025. 17(22).

3. Baudin, E., et al., Sunitinib for metastatic progressive phaeochromocytomas and paragangliomas: results from FIRSTMAPPP, an academic, multicentre, international, randomised, placebo-controlled, double-blind, phase 2 trial. Lancet, 2024. 403(10431): p. 1061–1070.

4. Jimenez, C., et al., Belzutifan for Advanced Pheochromocytoma or Paraganglioma. N Engl J Med, 2025. 393(20): p. 2012–2022.

5. Fojo, A.T., et al., Single agent axitinib in the management of patients with progressive pheochromocytoma and paraganglioma. Journal of Clinical Oncology, 2024. 42(16_suppl): p. 4602–4602.

6. Lin, F.I., et al., Phase II Study of (177)Lu-DOTATATE for Progressive Metastatic Pheochromocytomas and Paragangliomas: Interim Analysis of Efficacy, Safety, and Biomarkers. J Clin Oncol, 2025. 43(28): p. 3102–3112.

7. Belge Bilgin, G., et al., Outcomes of Peptide Receptor Radionuclide Therapy with (177)Lu-DOTATATE in Patients with Metastatic Pheochromocytoma and Paraganglioma. J Nucl Med, 2026. 67(1): p. 96–104.

8. Fischer, A., et al., Metastatic Pheochromocytoma and Paraganglioma: Somatostatin Receptor 2 Expression, Genetics, and Therapeutic Responses. J Clin Endocrinol Metab, 2023. 108(10): p. 2676–2685.

9. Patel, M., et al., Somatostatin Receptors and Analogs in Pheochromocytoma and Paraganglioma: Old Players in a New Precision Medicine World. Front Endocrinol (Lausanne), 2021. 12: p. 625312.

10. Leijon, H., et al., Variable somatostatin receptor subtype expression in 151 primary pheochromocytomas and paragangliomas. Hum Pathol, 2019. 86: p. 66–75.

11. Elston, M.S., et al., Increased SSTR2A and SSTR3 expression in succinate dehydrogenase-deficient pheochromocytomas and paragangliomas. Hum Pathol, 2015. 46(3): p. 390–6.

12. Hofman, M.S. and R.J. Hicks, Moving Beyond "Lumpology": PET/CT Imaging of Pheochromocytoma and Paraganglioma. Clin Cancer Res, 2015. 21(17): p. 3815–7.

13. Network, N.C.C., *NCCN Clinical Practice Guidelines in Oncology (NCCN Guidelines®)*: Neuroendocrine and Adrenal Tumors. 2026, National Comprehensive Cancer Network: Plymouth Meeting, PA.

14. Caplin, M.E., et al., Lanreotide in metastatic enteropancreatic neuroendocrine tumors. N Engl J Med, 2014. 371(3): p. 224–33.

15. Del Rivero, J., et al., Systemic Therapy for Tumor Control in Metastatic Well-Differentiated Gastroenteropancreatic Neuroendocrine Tumors: ASCO Guideline. J Clin Oncol, 2023. 41(32): p. 5049–5067.

16. Dromain, C., et al., Novel Tumor Growth Rate Analysis in the Randomized CLARINET Study Establishes the Efficacy of Lanreotide Depot/Autogel 120 mg with Prolonged Administration in Indolent Neuroendocrine Tumors. Oncologist, 2021. 26(4): p. e632–e638.

17. Nasca, V., et al., Sunitinib for the treatment of patients with advanced pheochromocytomas or paragangliomas: The phase 2 non-randomized SUTNET clinical trial. Eur J Cancer, 2024. 209: p. 114276.

18. Tang, C.Y.L., et al., Safety and efficacy of peptide receptor radionuclide therapy in patients with advanced pheochromocytoma and paraganglioma: A single-institution experience and review of the literature. J Neuroendocrinol, 2023. 35(12): p. e13349.

19. Tobimatsu, A., et al., Potentially fatal crisis after (177)Lu-DOTATATE therapy for paraganglioma: a case report with review of literature. Endocr J, 2025. 72(10): p. 1143–1150.

20. Fischer, A., et al., Responses to systemic therapy in metastatic pheochromocytoma/paraganglioma: a retrospective multicenter cohort study. Eur J Endocrinol, 2023. 189(5): p. 546–565.

